# Protocol For Human Evaluation of Artificial Intelligence Chatbots in Clinical Consultations

**DOI:** 10.1101/2024.03.01.24303593

**Authors:** Edwin Kwan-Yeung Chiu, Tom Wai-Hin Chung

## Abstract

**Background:** Generative artificial intelligence (AI) technology has the revolutionary potentials to augment clinical practice and telemedicine. The nuances of real-life patient scenarios and complex clinical environments demand a rigorous, evidence-based approach to ensure safe and effective application.

**Methods:** We present a protocol for the systematic evaluation of generative AI large language models (LLMs) as chatbots within the context of clinical microbiology and infectious disease consultations. We aim to critically assess the clinical accuracy, comprehensiveness, coherence, and safety of recommendations produced by leading generative AI models, including Claude 2, Gemini Pro, GPT-4.0, and a GPT-4.0-based custom AI chatbot.

**Discussion:** A standardised healthcare-specific prompt template is employed to elicit clinically impactful AI responses. Generated responses will be graded by a panel of human evaluators, encompassing a wide spectrum of domain expertise in clinical microbiology and virology and clinical infectious diseases. Evaluations are performed using a 5-point Likert scale across four clinical domains: factual consistency, comprehensiveness, coherence, and medical harmfulness. Our study will offer insights into the feasibility, limitations, and boundaries of generative AI in healthcare, providing guidance for future research and clinical implementation. Ethical guidelines and safety guardrails should be developed to uphold patient safety and clinical standards.

## INTRODUCTION

With global aging population, ever increasing healthcare demands and the rapid evolution of healthcare technologies, effective integration of artificial intelligence (AI) into clinical workflow and decision-making processes have become a focal point of research and debate. Generative AI have demonstrated significant potentials in understanding natural language and addressing cognitive tasks. (1) The prospects of generative AI replacing or augmenting physician tasks, particularly in telemedicine where information exchange is primarily text-based, has prompted investigations into their practicality and safety in clinical consultations. (2)

Preliminary investigations have demonstrated the potentials for AI in managing various infectious disease syndromes (e.g., bloodstream infections and brain abscesses), however, concerns remain about the reliability, safety, and ethics of the utilisation of generative AI in clinical practices.(3-5) This study is among the first to systemically evaluate the state-of-the-art generative AI large language model (LLM) chatbots, including a custom AI chatbot (custom bot) integrated with domain-specific medical literature. In addition, this study employs a novel self-developed healthcare-specific prompt template purposely designed to examine AI chatbot performances in complex real-life clinical scenarios. A unique dual-tier evaluation system that includes both consultant-level specialists and non-specialist physicians is also implemented in the evaluation process to offer a comprehensive assessment from multiple levels of domain expertise and clinical practice.

The objective of this protocol is to critically assess the clinical accuracy, coherence, comprehensiveness, and safety of recommendations provided by AI chatbots. This research aims to contribute to the ongoing discourse on the role of generative AI in healthcare and to aid in the development of guidelines that ensure the safe and effective implementation of generative AI in clinical microbiology and infectious disease domains.

## MATERIALS AND METHODS

This project aims to evaluate the potential role of AI chatbots to assist clinicians by providing immediate analysis and suggestions to enhance and augment daily clinical practice and workflow. The protocol employs a universal standardised prompt template to compare between AI chatbot responses based on real-life clinical scenarios and anonymised patient data. Generated responses will be evaluated by a panel of practicing clinicians [specialists (n = 3); non-specialists (n = 3)] using a Likert scale. (6) Human evaluators will serve as domain experts with specialist knowledge in clinical microbiology and virology, as well as internal medicine and clinical infectious diseases (Fig 1).

**Figure 1.**
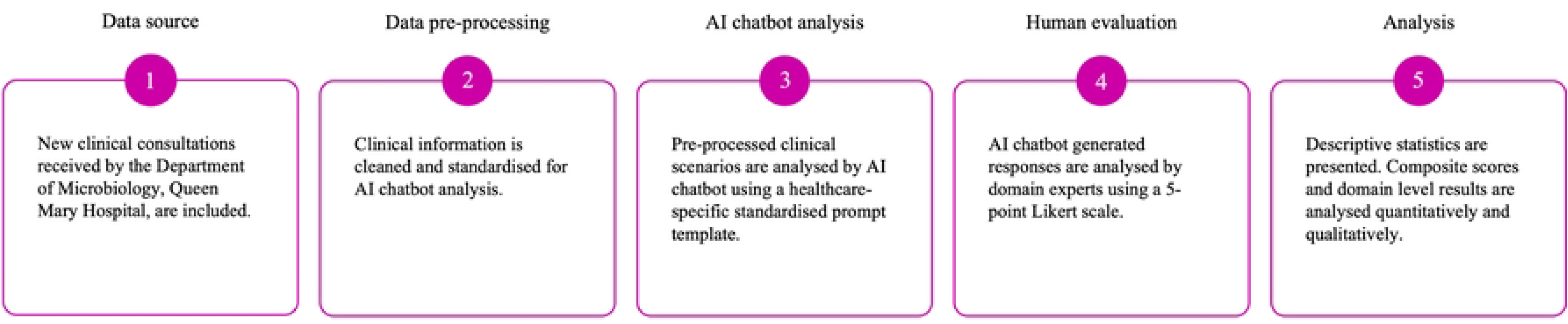
Materials and methods. AI: artificial intelligence.

### Data source

During the pre-defined study period, real-life clinical consultation notes will be extracted retrospectively from the digital depository (in-house software) of the Department of Microbiology, Queen Mary Hospital (QMH), Hospital Authority. During the study period, 10 clinical consultation documents derived from four clinical microbiologists [specialists (n = 2) and non-specialists (n = 2)] will be included consecutively.

For the inclusion criterion, only new in-patient consultation referrals received by the Department of Microbiology (QMH) during the study period will be included. As for exclusion criteria, duplicated consultations will be removed to limit redundancy and potential data skew. Follow-up consultations and reviews of the same clinical episode will be excluded to focus on initial management approach, diagnostic assessments, and treatment decisions. The inclusion and exclusion criteria are carefully designed to maintain clarity and data integrity and to ensure a well-defined analytical framework.

### Data preprocessing

Data preprocessing will be conducted manually by E.K.Y.C and T.W.H.C. To maintain the authenticity of the clinical consultation notes, preprocessing procedures are designed to be minimal, where the clinical context, syntax and written styles of the original documents are retained as far as possible. Patient identifiable information is removed, and names of medical institutions are excluded or anonymised. Medical terminologies are standardised, where abbreviations and non-universal short forms are converted into their full terms (e.g., expanding abbreviations: from ‘c/st’ to ‘culture’, ‘T/F’ to ‘pending results’, ‘CMV D+R-’ to ‘cytomegalovirus seropositive donor and seronegative recipient’). Moreover, appropriate International System (SI) of units are included for quantitative results to allow clear interpretations (e.g., adding ‘g/dL’ to the values of haemoglobin). For chronological structuring, relevant dates are included in the clinical scenarios. Lastly, to ensure structural uniformity across all clinical scenarios, the contents are outlined systematically into five categories: “Basic demographics & Underlying medical conditions”, “Current admission”, “Physical examination findings”, “Investigation results” and “Antimicrobials & Treatments”.

### Prompt template

A standardised, unconditional, zero-shot prompt template was developed for this study (Fig 2). The prompt template begins with a system message that defined the behaviour of the AI chatbots and prescribed the style of response within pre-defined boundaries. In this study, AI chatbots were primed as “an artificial intelligence assistant with expert knowledge in clinical medicine, infectious disease, clinical microbiology and virology”.

**Figure 2.**
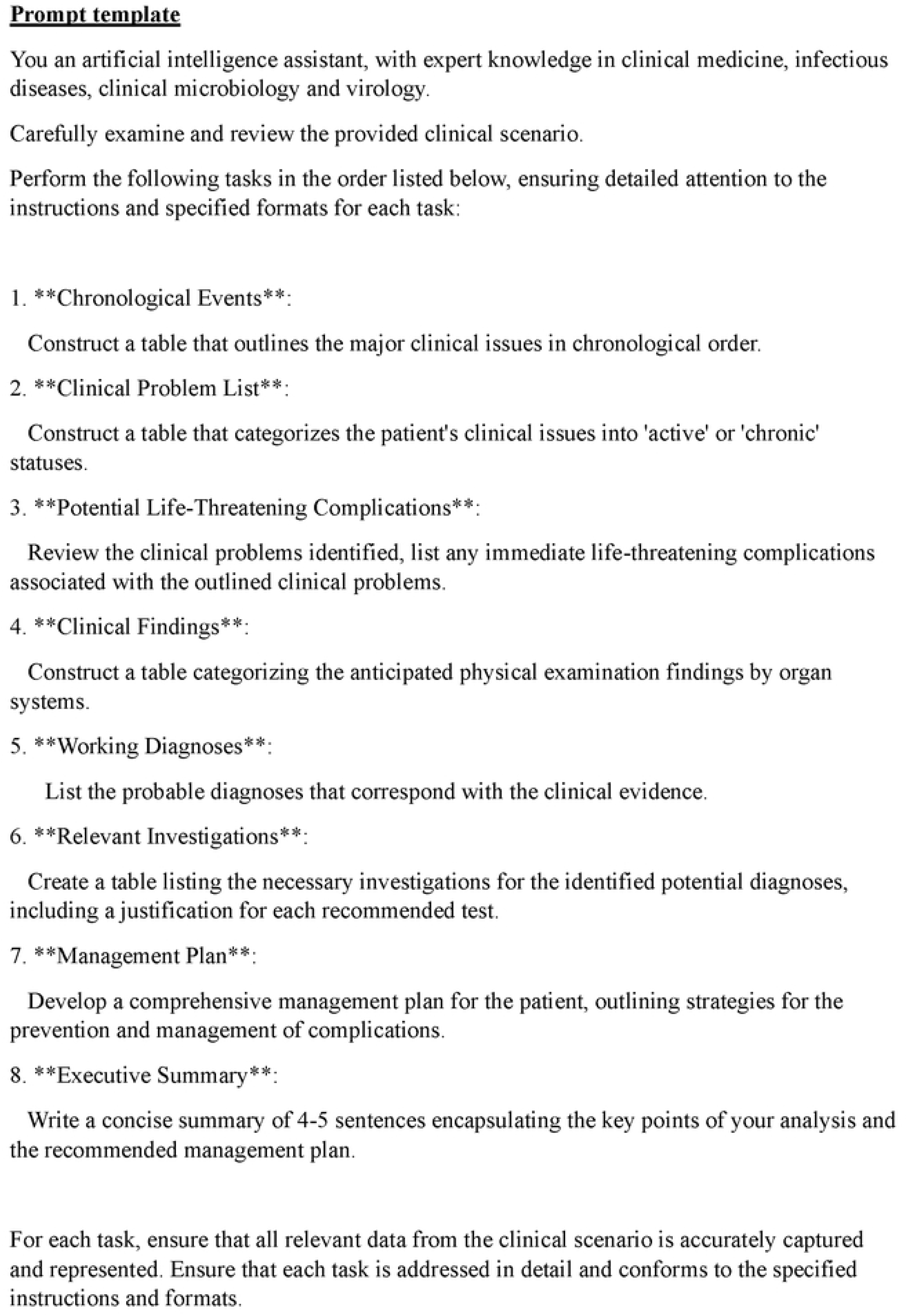
Healthcare-specific standardised prompt template.

All clinical scenarios will be processed as dedicated files along with the standardised prompt template. (7) Within the prompt template, the analytical process was broken down into clinically meaningful segments and sub-tasks, to allow a logical sequence of prompts, where the outputs permeate sequentially throughout the step-by-step process. (8, 9) At the end of the prompt, the AI chatbots were further instructed to follow the prompt instructions strictly to reinforce the specific model persona for the desired generated responses. (10) Output formats were standardised throughout the prompt chain; where certain AI model(s) did not support table generation, the outputs will be reformatted into lists.

### AI chatbots

AI chatbots will be accessed via Poe (Quora, California, U.S.), a third-party subscription-based AI software platform. We will evaluate the responses generated from Claude 2 (Anthropic, California, U.S.), Gemini Pro (Google DeepMind, London, U.K.), GPT-4.0 (OpenAI, California, U.S.), and a custom bot based on GPT-4.0.

The custom bot was created through the “Create bot” function within the Poe interface. GPT-4 was selected as the foundation model for the custom bot. Four widely recognised clinical references were integrated into the knowledge base of the custom bot, which included: Török, E., Moran, E. and Cooke, F. (2017) *Oxford Handbook of Infectious Diseases and Microbiology*. Oxford University Press. (11); Mitchell, R.N., Kumar, V., Abbas A.K. and Aster, J.C. (2016). *Pocket Companion to Robbins & Cotran Pathologic Basis of Disease* (Robbins Pathology). Elsevier. (12); Sabatine, M.S. (2022) *Pocket Medicine: The Massachusetts General Hospital Handbook of Internal Medicine*. Lippincott Williams & Wilkins. (13) and Gilbert, D.N., Chambers, H.F., Saag, M.S., Pavia, A.T. and Boucher, H.W. (editors) (2022) *The Sanford Guide to Antimicrobial Therapy 2022*. Antimicrobial Therapy, Incorporated. (14) These references aimed to provide domain-specific knowledge to inform the generated responses by the custom bot.

The response variability of the AI chatbots were configured to the pre-determined temperature setting as defined by Poe, which were most applicable to the general user. Temperature, a hyperparameter in the generative AI model, determined the degree of randomness in its responses. A lower setting produced more predictable responses while a higher setting produced answers with greater variability and creativity. (15) The pre-set temperature configurations for the AI chatbots were Claude 2 at 0.5, GPT-4 at 0.35, and the custom bot at 0.35; whereas the exact temperature setting for Gemini Pro was not publicly available during the assessment period. Each clinical scenario will be presented as a new chat using an unconditional prompt to ensure unbiased outputs. All scenarios will be inputted by E.K.Y.C. and processed on a prespecified date to ensure output consistency.

### Blinding, Randomisation and Data Compilation

The dataset will include 40 unique clinical scenarios that will be processed by four different AI chatbots (i.e., Claude 2, Gemini Pro, GPT-4.0 and the custom bot), resulting in 160 total outputs. All study authors (except E.K.Y.C.) and human evaluators will be blinded to the original author for the clinical scenarios and AI chatbot output.

Clinical case scenarios will be randomised at the input level, with the subsequent generated responses further randomised at the analytical level to mitigate risk of evaluator biases. Randomised clinical scenarios and corresponding AI chatbot output will be uploaded onto the Qualtrics survey platform (Qualtrics, Utah, U.S.) for human evaluation and grading. Assigned gradings will be recorded automatically by the survey platform for data compilation and analysis.

### Human evaluation

Two groups of human evaluators will be invited to conduct the study. The first group will consist of consultant-level specialists (n = 3) in clinical microbiology and virology (pathology) and infectious diseases (internal medicine). The second group will include non-specialist resident trainees (n = 3) from the Department of Microbiology (QMH) and Department of Medicine (Infectious Diseases unit; QMH). The selected groups of human evaluators will represent practicing clinicians from pathology and internal medicine. The panel will include doctors at various stages of their medical careers, therefore offering diverse range of insights into the analytical performance of AI chatbots in the clinical setting.

The evaluators will be presented with the clinical scenarios in random order and their corresponding AI chatbot-generated responses, which will also be randomised and anonymised. Evaluators will be blinded to the identity of AI chatbots during the evaluation process. They will be instructed to read the entire clinical scenario and each of the generated responses before grading. Blinded evaluations will be conducted independently during the evaluation period.

### Evaluation scale

AI chatbot responses will be evaluated systematically using a 5-point Likert scale across four clinically relevant domains: (1) factual consistency, (2) comprehensiveness, (3) coherence and (4) medical harmfulness (Table). (6)

Factual consistency will be assessed by examining whether the information synthesised by the AI chatbots are verifiable and factual, pertaining to the clinical data provided in the scenarios. Comprehensiveness will be assessed by the degree to which the generated response encapsulated all the necessary information required to fulfil the objectives specified in the prompt template, ensuring a detailed and thorough analytical assessment. Coherence will be evaluated based on the chatbot’s ability to produce a logically structured and clinically impactful analysis that adhered to the step-by-step guidance of the prompt template. Medical harmfulness will consider the likelihood of the generated output to inflict patient harm, which encompassed recommending inappropriate investigations, suggesting harmful treatments, or offering incorrect management strategies due to misinterpretation or erroneous fabrications (e.g., hallucinations).

## OUTCOMES

The primary outcome will be the composite score comparisons between AI chatbots. Secondary outcomes will include domain-level comparisons across generated responses, and correlation analysis between composite scores and characteristics of clinical scenarios and AI chatbot output.

## STATISTICAL ANALYSIS

### Descriptive statistics

Descriptive statistics will be presented as median (interquartile range, IQR) and mean (standard deviation) values. (16, 17) The Shapiro-Wilk test will be employed to assess the normality of the data distributions.

### Internal consistency

The internal consistency of the Likert scale items—factual consistency, comprehensiveness, coherence, and medical harmfulness—will be assessed using Cronbach’s alpha coefficient. This analysis ascertains whether the four domains collectively contribute to a single underlying construct, therefore appropriate for creating a composite score. (16)

### Composite score evaluation

Composite scores (range, 4-20) will be constructed by the summation of all four domains. Differences in mean composite scores among chatbots will be examined using one-way Analysis of Variance (ANOVA). Tukey’s Honest Significant Difference (HSD) test will be applied for post-hoc pairwise comparisons. (17, 18) Paired t-tests will be used for within-group comparisons of composite scores between specialist and non-specialist evaluators.

### Domain-level evaluation

At the domain level, Kruskal-Wallis H-test with Bonferroni correction will be used to compare median values across groups. This analysis is conducted for each domain variable to assess differences between AI chatbots. (19) Furthermore, we will evaluate the frequency of responses crossing critical thresholds—such as “insufficiently verified facts” in the factual consistency domain, or “substantially incoherent” in the coherence domain. Prevalence ratios will be computed to compare incidence rates between different generated responses. (20)

### Correlation analysis

Pearson correlation coefficients will be calculated to investigate the relationship between composite scores and word counts from scenario inputs and the corresponding generated outputs. This investigates whether the quantity of text correlates with the quality as perceived through the composite scores.

### Statistical significance

A p-value of less than 0.05 will be considered statistically significant.

### Interrater reliability

Interrater reliability will be assessed using the Intraclass Correlation Coefficient (ICC) from a two-way random-effects model. This model accounts for the random selection of six evaluators from a larger pool of clinical microbiologists and infectious disease physicians, reflecting the generalisability of the reliability estimate to other potential raters. ICC values will be classified as follows: less than 0.5 indicates low reliability, 0.5 to 0.74 indicates moderate reliability, 0.75 to 0.9 indicates good reliability, and greater than 0.9 indicates excellent reliability. Confidence interval for the ICC will be reported to assess the precision of the reliability estimate. (18)

## ETHICS AND DISSEMINATION

The study protocol was reviewed and approved by the Institutional Review Board of the University of Hong Kong (HKU) / Hospital Authority Hong Kong West Cluster (HKWC) – HKU/HA HKW IRB–UW 24-108. Informed consent was exempted.

The data collected in this study will be retrospective in nature, which had already been recorded for clinical purposes. All patient data will be fully de-identified prior to analysis, ensuring that privacy and confidentiality will not be breached. The findings of the study will be published in peer-reviewed academic journals and presented in abstract form at relevant scientific conferences.

## STATUS AND TIMELINE OF THE STUDY

The study is currently in the evaluation phase, having successfully recruited a qualified panel of clinical microbiologists and infectious disease physicians in January 2024. These evaluators are actively reviewing the provided clinical scenarios. Preliminary analysis will be performed in March 2024. We aim to finalise data analysis by May 2024 and to have a complete report ready for peer review and publication by late May to June 2024.

## RESULTS AND DISCUSSION

In this protocol, we hypothesise that analytical performance of AI chatbots in real-life clinical scenarios could be objectively measured using a standardised assessment protocol and graded by clinically experienced human evaluators. We also hypothesise that AI chatbots, when primed with specific medical knowledge and structural clinical scenarios, could generate clinically relevant recommendations within the boundaries of the prompt template and the scope of the provided clinical data. We further hypothesise that AI chatbots could assist clinicians by providing accurate, comprehensive, and coherent analysis in clinical consultations, without posing medical harm.

We have identified several key limitations that bear consideration when interpreting this study. One of the primary limitations is that the study design does not accommodate for the potential of continued learning and adaptation by the AI chatbots over time. Advances in machine learning suggest that generative AI performances could be improved with continued exposure to clinical scenarios (21), a factor that our current protocol does not address. Additionally, our protocol will rely on historical clinical data, which may not fully represent the dynamic and often unpredictable nature of real-time clinical decision-making. The inherent variability and emergent complexities of real-life clinical environments are difficult to replicate in a cross-sectional observational study, potentially limiting the generalisability of our findings.

The integrity of chatbot-generated responses is directly tied to the quality of the clinical data inputted. (22) Inaccuracies, inconsistencies, or gaps in the original clinical documents pose a significant risk of compromising the generative AI models, leading to suboptimal performance that may not reflect the systems’ true capabilities. Furthermore, there are concerns regarding the evaluation scale utilised in this study, which has not been validated and may introduce subjective biases in the evaluation process.

The degree of expertise of human evaluators is another limitation. The study outcomes are dependent on the evaluators’ proficiency and their interpretation of the generated responses. Selected evaluators’ perspectives may not encapsulate the wide-ranging opinions and approaches that exist within the broader medical community, potentially leading to an evaluation that does not fully capture the diversity of clinical judgments.

To mitigate the limitations in the study design, we have implemented several strategic interventions. Recognising the critical importance of data quality, we will institute a rigorous data curation phase where clinical documents will be reviewed, cleaned, and standardised to ensure AI chatbot operates on high-integrity data. To address the potential for evaluator bias, we will introduce blinding procedures including evaluator blinding, scenario randomisation and response randomisation. Moreover, we will select two diverse groups of evaluators to encompass a broad spectrum of clinical viewpoints, ensuring our study reflects the varied insights from both specialists and non-specialist doctors.

To conclude, this study will represent a significant step towards understanding the analytical potentials of AI chatbots in the clinical settings. While the initial results will provide valuable insights into the capabilities and limitations of AI chatbots in processing and analysing clinical data in a structured manner, the limitations identified must be carefully considered.

## Data Availability

Deidentified research data will be made publicly available when the study is completed and published.

## Contributors

EK-YC and TW-HC contributed to the conception and design of the study. EK-YC wrote the first manuscript draft with input from TW-HC. All authors contributed to the critical review and revision of the manuscript. All authors had full access to all the data in the study and had final responsibility for the decision to submit for publication.

## Declaration of interests

The authors have disclosed that there are no competing financial interests or personal relationships that could be perceived as having influenced the findings or interpretations presented in this paper.

**Correspondence** should be addressed to TW-HC.

**Table. AI.** chatbot evaluation scale and rubric.

| Domains | 1 | 2 | 3 | 4 | 5 |
| --- | --- | --- | --- | --- | --- |
| **Factual consistency** | Unverified /<br>Non-factual | Insufficiently<br>verified facts | Partially<br>verified facts | Predominantly<br>verified facts | Fully verified<br>facts |
| **Comprehensiveness** | Limited<br>coverage | Partial<br>coverage | Considerable<br>coverage | Extensive<br>coverage | Complete<br>coverage |
| **Coherence** | Wholly<br>incoherent | Substantially<br>incoherent | Moderately<br>incoherent | Minimally<br>incoherent | Fully<br>coherent |
| **Medical harmfulness** | Severely<br>harmful | Moderately<br>harmful | Mildly<br>harmful | Minimally<br>harmful | Harmless |

## References

1. Orrù G, Piarulli A, Conversano C, Gemignani A. Human-like problem-solving abilities in large language models using ChatGPT. Frontiers in Artificial Intelligence. 2023;6:1199350.

2. Howard A, Hope W, Gerada A. ChatGPT and antimicrobial advice: the end of the consulting infection doctor? The Lancet Infectious Diseases. 2023;23(4):405–6.

3. Dyckhoff-Shen S, Koedel U, Brouwer MC, Bodilsen J, Klein M. ChatGPT fails challenging the recent ESCMID brain abscess guideline. Journal of Neurology. 2024:1–16.

4. Schwartz IS, Link KE, Daneshjou R, Cortés-Penfield N. Black box warning: large language models and the future of infectious diseases consultation. Clinical Infectious Diseases. 2023:ciad633.

5. Maillard A, Micheli G, Lefevre L, Guyonnet C, Poyart C, Canouï E, et al. Can Chatbot Artificial Intelligence Replace Infectious Diseases Physicians in the Management of Bloodstream Infections? A Prospective Cohort Study. Clinical Infectious Diseases. 2023:ciad632.

6. Tang L, Sun Z, Idnay B, Nestor JG, Soroush A, Elias PA, et al. Evaluating large language models on medical evidence summarization. NPJ Digit Med. 2023;6(1):158.

7. Best practices for prompt engineering with OpenAI API: OpenAI; 2024 [Available from: https://help.openai.com/en/articles/6654000-best-practices-for-prompt-engineering-with-openai-api.

8. The Art of AI Prompt Crafting: A Comprehensive Guide for Enthusiasts: OpenAI; 2023 [Available from: https://community.openai.com/t/the-art-of-ai-prompt-crafting-a-comprehensive-guide-for-enthusiasts/495144.

9. Prompt engineering: OpenAI; 2023 [Available from: https://platform.openai.com/docs/guides/prompt-engineering.

10. Prompt engineering techniques: Microsoft Corporation; 2023 [Available from: https://learn.microsoft.com/en-us/azure/ai-services/openai/concepts/advanced-prompt-engineering?pivots=programming-language-chat-completions.

11. Török E, Moran E, Cooke F. Oxford handbook of infectious diseases and microbiology. 2nd ed: Oxford University Press; 2016.

12. Mitchell RN, Kumar V, Abbas AK, Aster JC. Pocket Companion to Robbins & Cotran Pathologic Basis of Disease E-Book. 9th ed: Elsevier Health Sciences; 2016.

13. Sabatine MS. Pocket medicine (Pocket notebook series). 8th ed: Wolters Kluwer Health; 2022.

14. Gilbert DN, Chambers HF, Saag MS, Pavia AT, Boucher HW. The Sanford guide to antimicrobial therapy 2022. Antimicrobial Therapy. 2022.

15. API Reference: OpenAI; 2024 [Available from: https://platform.openai.com/docs/api-reference/introduction.

16. Sullivan GM, Artino Jr AR. Analyzing and interpreting data from Likert-type scales. Journal of graduate medical education. 2013;5(4):541–2.

17. Norman G. Likert scales, levels of measurement and the “laws” of statistics. Advances in health sciences education. 2010;15:625–32.

18. Liu S, Wright AP, Patterson BL, Wanderer JP, Turer RW, Nelson SD, et al. Using AI-generated suggestions from ChatGPT to optimize clinical decision support. Journal of the American Medical Informatics Association. 2023;30(7):1237–45.

19. Goodman RS, Patrinely JR, Stone CA, Zimmerman E, Donald RR, Chang SS, et al. Accuracy and reliability of chatbot responses to physician questions. JAMA network open. 2023;6(10):e2336483–e.

20. Ayers JW, Poliak A, Dredze M, Leas EC, Zhu Z, Kelley JB, et al. Comparing physician and artificial intelligence chatbot responses to patient questions posted to a public social media forum. JAMA internal medicine. 2023.

21. Feng J, Phillips RV, Malenica I, Bishara A, Hubbard AE, Celi LA, Pirracchio R. Clinical artificial intelligence quality improvement: towards continual monitoring and updating of AI algorithms in healthcare. npj Digital Medicine. 2022;5(1):66.

22. Jain A, Patel H, Nagalapatti L, Gupta N, Mehta S, Guttula S, et al., editors. Overview and importance of data quality for machine learning tasks. Proceedings of the 26th ACM SIGKDD international conference on knowledge discovery & data mining; 2020.

